# The Dearth of Representation in FDA Approved Drug Trials

**DOI:** 10.1101/2024.01.16.24301376

**Authors:** Christine Ibilibor, Shannon Armbruster, Rell Parker, Jia-Ray Yu, Andrew Barros

**Author notes:** Corresponding author: Andrew Barros MD.

## Abstract

The generalizability of data derived from randomized controlled trials is of paramount importance given their utility in the Food & Drug Administration (FDA) drug approval process. An essential part of this process is the inclusion of reliably reported gender, race and ethnicity data in trials that lead to FDA drug approval. Despite previous mandates by the FDA and Clinicaltrials.gov, gender and race-specific data remains under reported. We reviewed 100 most recently approved FDA medications, and abstracted the clinical trial data from Clinicaltrials.gov that supported their approval. We then compared these FDA approved trials to non-FDA approved trials from the same year and of similar size. We found that 40% of the FDA trials were missing race/ethnicity information, while 24% of these trials did not include gender information. We demonstrate that there remains a significant amount of missing gender and racial/ethnic data in trials that lead to FDA-approved medications.

## Introduction

In 2016, the Food and Drug Administration (FDA) set expectations for the use of a standardized approach for collecting data, using standard terminology for age, sex, gender, race, and ethnicity^1–3^. Subsequently, in 2017 ClinicalTrials.gov instituted the requirement of reporting of race/ethnicity information (if collected) during results submission for trials. In one study including data from ClinicalTrials.gov from March 2000 to March 2020, gender information was available for 19,866 of 20,020) of studies (99.2%)^4^. Disparities in female participants were apparent, being underrepresented in several fields relative to their disease burden^4^. During that same time period, race and ethnicity reporting increased from 42% to 91.4%, but many trials (22.9%) still did not use the classifications as recommend by the NIH/US Office of Budget and Management (NIH OMB)^2^. Given the high level of reporting in FDA-approved drug trials and the importance of diverse representation within these trials, we sought to investigate racial, ethnic, and gender-specific representation in FDA approved drug clinical trials.

## Methods

We reviewed FDA drug announcements for the 100 most recently approved medications (excluding radiopharmacuticals and medications approved through non-traditional pathways) and abstracted the clinical trial data that supported their approval (FDA trials). For each trial, we used an Aggregate Content of ClinicalTrials.gov (https://aact.ctti-clinicaltrials.org), a publicly available relation database extract of ClinicalTrials.gov, to identify 5 interventional, randomized trials from the same year and closest in size to our case trial (Control trials). For all trials, we extracted the total population size, number of female participants, and number of white participants. We evaluated the proportion of participants using generalized additive logistic regression model with a spline term for trial size. We have followed the EQUATOR guidelines.

## Results

In total, we identified 107 trials supporting FDA-approved drugs and 535 control trials (Table 1). Forty percent of the FDA trials were missing race/ethnicity information, while 24% of trials did not include gender information. When compared to control studies, trials supporting drug submissions were significantly more likely to report gender (OR 8.30, 95% CI 5.2 – 13.65) and race (OR 10.05, 95% CI 6.37 – 16.06). Figure 1 shows that FDA trials had an average marginal effect of including 7.29% more women (95% CI 6.77% - 7.82%) and 10.14% more white participants (95% CI 9.56% - 10.73%).

| Characteristic | Control Trials, N =<br>535<sup>1</sup> | FDA Trials, N =<br>107<sup>1</sup> | p-value<sup>2</sup> |
| --- | --- | --- | --- |
| Year of Trial Completion |  |  | >0.9 |
| - 2011 | 25 (4.7%) | 5 (4.7%) |  |
| 2011 - 2015 | 75 (14%) | 15 (14%) |  |
| 2016 - 2020 | 370 (69%) | 74 (69%) |  |
| 2021 - | 65 (12%) | 13 (12%) |  |
| Total Participants | 358 (156,<br>731) | 358 (162,<br>728) | >0.9 |
| Female Participants, % | 0.52 (0.39,<br>0.63) | 0.62 (0.48,<br>0.83) | 0.002 |
| Gender Not Reported to<br>ClinicalTrials.gov | 389 (73%) | 26 (24%) | <0.001 |
| White Participants, % | 0.73 (0.51,<br>0.86) | 0.78 (0.65,<br>0.91) | 0.070 |

| Characteristic | Control Trials, N = 535<sup>1</sup> | FDA Trials, N = 107<sup>1</sup> | p-value<sup>2</sup> |
| --- | --- | --- | --- |
| Race Not Reported to ClinicalTrials.gov | 466 (87%) | 43 (40%) | <0.001 |

<sup>1</sup> n (%); Median (IQR)
<sup>2</sup> Fisher's exact test; Wilcoxon rank sum test; Pearson's Chi-squared test

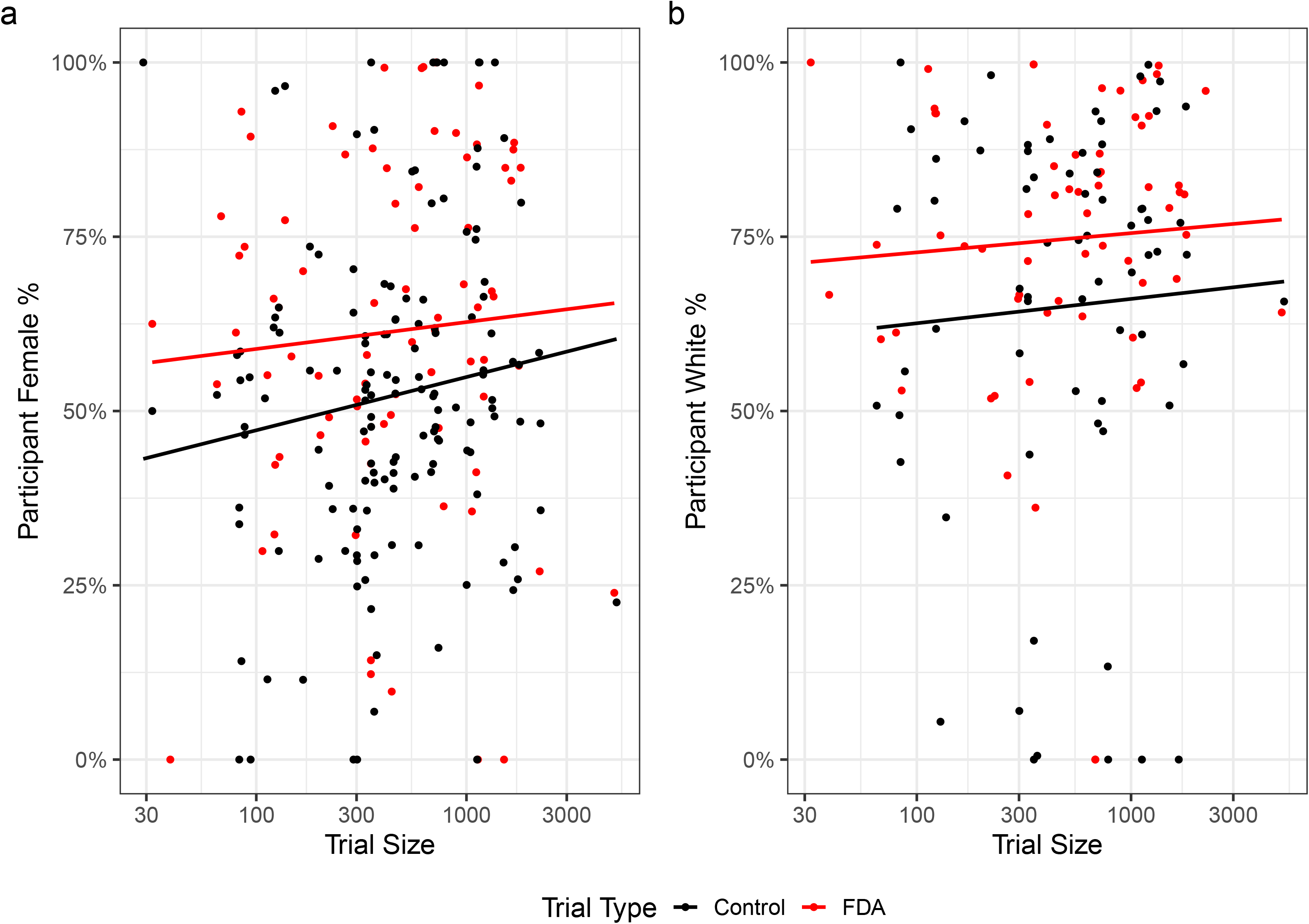

## Discussion

We demonstrate that there remains a significant amount of missing gender and racial/ethnic data in trials that lead to FDA-approved medications. Gender and racial diversity in clinical trials, particularly those that lead to practice defining FDA approved agents, is important for data generalizability. The FDA mandate has improved the rates of reporting gender and race as shown in our study in which FDA-approved trials were 10 times more likely to report gender and race compared to non-FDA-approved trials.

One limitation of our study is that we primarily utilized the NIH OMB classification for race which reduced our sample to studies using this nomenclature. The NIH OMB racial/ethnic classification, while widely used, might not capture the breadth of cultural differences within a race and across races that are clinically meaningful. Recognition of the wide range of racial, ethnic, and gender-based diversity among participants involved in research has become of increasing importance in the scientific community since these classifications were first implemented. The upcoming update to the NIH OMB racial classification system is a reflection of the changing social and cultural landscape that will allow for capturing more granular data on diversity in clinical research^5^. Additionally, some trials may have not included sex or gender information if the trial was completed in a population that only includes one sex (ex. Males and prostate cancer, or females are ovarian cancer).

## Data Sharing Statement

The source code to support this analysis is available at https://github.com/ajb5d/RaceAndGenderInClinicalTrials. The data will be accessible by accessing the source code through GitHub and the iTHRIV data commons.

## Acknowledgements

All authors are members of the iTHRIV Scholars program. The iTHRIV Scholars Program is supported in part by the NCATS of the NIH under Award Numbers UL1TR003015 and KL2TR003016.

## Author Contributions

All authors had full access to all of the data in the study and take responsibility for the integrity of the data and the accuracy of the data analysis.

Concept and design: All authors.

Acquisition, analysis, or interpretation of data: All authors.

Drafting of the manuscript: All authors.

Critical review of the manuscript for important intellectual content: All authors.

Statistical analysis: Andrew Barros

Administrative, technical, or material support: All authors.

